# Evidence for and level of herd immunity against SARS-CoV-2 infection: the ten-community study

**DOI:** 10.1101/2020.09.24.20200543

**Authors:** Andrew Jeremijenko, Hiam Chemaitelly, Houssein H. Ayoub, Moza Abdellatif Hassan Abdulla, Abdul-Badi Abou-Samra, Jameela Ali A.A. Al Ajmi, Nasser Ali Asad Al Ansari, Zaina Al Kanaani, Abdullatif Al Khal, Einas Al Kuwari, Ahmed Al-Mohammed, Naema Hassan Abdulla Al Molawi, Huda Mohamad Al Naomi, Adeel A. Butt, Peter Coyle, Reham Awni El Kahlout, Imtiaz Gillani, Anvar Hassan Kaleeckal, Naseer Ahmad Masoodi, Anil George Thomas, Hanaa Nafady-Hego, Ali Nizar Latif, Riyazuddin Mohammad Shaik, Nourah B M Younes, Hanan F. Abdul Rahim, Hadi M. Yassine, Mohamed G. Al Kuwari, Hamad Eid Al Romaihi, Sheikh Mohammad Al Thani, Roberto Bertollini, Laith J. Abu-Raddad

**Affiliations:** Hamad Medical Corporation, Doha, Qatar; Infectious Disease Epidemiology Group, Weill Cornell Medicine-Qatar, Cornell University, Doha, Qatar; World Health Organization Collaborating Centre for Disease Epidemiology Analytics on HIV/AIDS, Sexually Transmitted Infections, and Viral Hepatitis, Weill Cornell Medicine–Qatar, Cornell University, Qatar Foundation – Education City, Doha, Qatar; Department of Mathematics, Statistics, and Physics, Qatar University, Doha, Qatar; Microbiology and Immunology Department, Faculty of Medicine, Assiut University, Assiut, Egypt; College of Health Sciences, QU Health, Qatar University, Doha, Qatar; Biomedical Research Center, Qatar University, Doha, Qatar; Department of Biomedical Science, College of Health Sciences, Member of QU Health, Qatar University, Doha, Qatar; Primary Health Care Corporation, Doha, Qatar; Ministry of Public Health, Doha, Qatar; Department of Population Health Sciences, Weill Cornell Medicine, Cornell University, New York, New York, USA

**Keywords:** SARS-CoV-2, epidemiology, COVID-19, infection, seroprevalence, immunity

## Abstract

**Background:** Qatar experienced a large severe acute respiratory syndrome coronavirus 2 (SARS-CoV-2) epidemic that disproportionately affected the craft and manual workers (CMWs) who constitute 60% of the population. This study aimed to investigate level of immunity in communities within this population as well as infection exposure required to achieve herd immunity.

**Methods:** Anti**-**SARS-CoV-2 seropositivity was assessed in ten CMW communities between June 21 and September 9, 2020. PCR positivity, infection positivity (antibody and/or PCR positive), and infection severity rate were also estimated. Associations with anti-SARS-CoV-2 positivity were investigated using regression analyses.

**Results:** Study included 4,970 CMWs who were mostly men (95.0%) and <40 years of age (71.5%). Seropositivity ranged from 54.9% (95% CI: 50.2-59.4%) to 83.8% (95% CI: 79.1-87.7%) in the different CMW communities. Pooled mean seropositivity across all communities was 66.1% (95% CI: 61.5-70.6%). PCR positivity ranged from 0.0% to 10.5% (95% CI: 7.4-14.8%) in the different CMW communities. Pooled mean PCR positivity was 3.9% (95% CI: 1.6-6.9%). Median cycle threshold (Ct) value was 34.0 (range: 15.8-37.4)—majority (79.5%) of PCR-positive individuals had Ct value >30 indicative of earlier rather than recent infection. Infection positivity (antibody and/or PCR positive) ranged from 62.5% (95% CI: 58.3-66.7%) to 83.8% (95% CI: 79.1-87.7%) in the different CMW communities. Pooled mean infection positivity was 69.5% (95% CI: 62.8-75.9%). Only five infections were ever severe and one was ever critical—an infection severity rate of 0.2% (95% CI: 0.1-0.4%).

**Conclusions:** Based on an extended range of epidemiological measures, active infection is rare in these communities with limited if any sustainable infection transmission for clusters to occur. At least some CMW communities in Qatar have reached or nearly reached herd immunity for SARS-CoV-2 infection at a proportion of ever infection of 65-70%.

## Introduction

Since the start of the severe acute respiratory syndrome coronavirus 2 (SARS-CoV-2) epidemic, millions of infections have been laboratory-confirmed globally [1], and millions others must have gone undocumented [2]. Two key questions remain unanswered: has any community reached herd immunity to render infection transmission chains unsustainable? What level of exposure to the infection (attack rate) is needed to reach herd immunity?

Qatar, a peninsula in the Arabian Gulf with a diverse population of 2.8 million [3], experienced a large-scale SARS-CoV-2 epidemic [4, 5]. By August 27, 2020, the rate of laboratory-confirmed infections in Qatar was at 50,324 per million population, one of the highest worldwide [6, 7]. The epidemic, currently in an advanced stage [4], seems to have followed a classic susceptible-infected-recovered “SIR” pattern with an epidemic peak around May 20 followed by a steady decline for the next four months [4].

The most affected subpopulation by this epidemic was that of the expatriate craft and manual workers (CMWs) among whom community transmission was first identified [4]. These workers constitute about 60% of the Qatar population and are typically single men aged 20-49 years [8]. CMWs at a given workplace or company not only work together during the day, but also live together as a *community* in large dormitories or housing complexes where they share rooms, bathrooms, and cafeteria-style meals [4, 9, 10]. These communities stay mostly in contact with their own community members and infrequently mingle with other communities, creating a geographic “bubble” that proved important for the pattern of infection transmission [4]. With reduced options for effective social and physical distancing, SARS-CoV-2 transmission in these CMW communities resembled that of influenza outbreaks in schools [4, 11, 12], and especially boarding schools [12].

Given the large number of diagnosed infections in CMWs [4], the large proportion of infections that were asymptomatic [4], the high polymerase chain reaction (PCR)-positivity rates in the random testing campaigns conducted around the epidemic peak in different CMW communities [4], and the observed “SIR” epidemic curve with rapid declines in incidence for over four months despite easing of the social and physical distancing restrictions [4], all pose a question as to whether herd immunity may have been reached in at least some of these communities.

Our aim was to assess ever exposure to the SARS-CoV-2 infection and attainment of herd immunity in several CMW communities by assessing the level of detectable antibodies. Operationally, we defined herd immunity as the proportion of the population ever infected (“attack rate”) beyond which infection transmission/circulation becomes unsustainable in this population with limited if any new infections occurring. The study was conducted to inform the national response and preparedness for potential future infection waves.

## Methods

### Data sources

Testing for detectable SARS-CoV-2-specific antibodies on blood specimens was conducted in ten CMW communities between June 21 and September 9, 2020 as part of a priori designed study combined with a testing and surveillance program led by the Ministry of Public Health (MOPH) and Hamad Medical Corporation (HMC), the main public healthcare provider and the nationally-designated provider for all COVID-19 healthcare needs in Qatar. The goal of this program was to assess the level of infection exposure in different subpopulations and economic sectors.

Study design was opportunistic utilizing the MOPH-HMC program and the need for rapid data collection to inform the national response. The ten CMW communities were selected for feasibility and/or given earlier random PCR testing campaigns or contact tracing that suggested substantial infection levels. For instance, CMW Community 1 was part of a random PCR testing campaign that identified a high positivity rate of 59% in late April. In six select communities, PCR testing was also simultaneously conducted to assess active infection using nasopharyngeal swabs, and in one select community (CMW Community 1), an interview schedule (based on World Health Organization (WHO) suggested questionnaire [13]) was administered to collect data on socio-demographics and history of exposure and symptoms.

The population size of each of these communities ranged from few hundreds to few thousands who live in shared accommodations provided by the employers. The companies that employ these workers belonged to the service or industrial sectors, but the bulk of the employees, even in the industrial companies, worked on providing services such as catering, cleaning and other janitorial services, warehousing, security, and port workers.

Employers were contacted and those agreeing to participate were asked to advertise the availability and location of testing sites to their employees. Individuals’ participation was voluntary. Employees interested in being tested and in knowing their status were provided with transportation to HMC testing sites. Informed consent and questionnaire were provided and collected in nine languages (Arabic, Bengali, English, Hindi, Urdu, Nepali, Sinhala, Tagalog, and Tamil) to cater to the main language groups in these CMW communities. National guidelines and standard of care were applied to all identified PCR positive cases. No action was mandated by the national guidelines to those found antibody positive, and thus no action was taken apart from notifying individuals of their sero-status.

Results of the serological testing were subsequently linked to the HMC centralized and standardized database comprising all SARS-CoV-2 PCR testing conducted in Qatar since the start of the epidemic [4, 6]. The database also includes data on hospitalization and on the WHO severity classification [14] for each laboratory-confirmed infection.

The study was approved by HMC and Weill Cornell Medicine-Qatar Institutional Review Boards.

### Laboratory methods

Testing for SARS-CoV-2-specific antibodies in the serological samples was performed using an electrochemiluminescence immunoassay, the Roche Elecsys^®^ Anti-SARS-CoV-2 (Roche, Switzerland). Results’ interpretation was per manufacturer’s instructions: reactive for cutoff index ≥ 1.0 and non-reactive for cutoff index <1.0 [15].

PCR testing was performed on aliquots of Universal Transport Medium (UTM) used for nasopharyngeal swabs’ collection (Huachenyang Technology, China). Aliquots were: extracted on the QIAsymphony platform (QIAGEN, USA) and tested with real-time reverse-transcription PCR (RT-qPCR) using the TaqPath™ COVID-19 Combo Kit (Thermo Fisher Scientific, USA) on a ABI 7500 FAST (ThermoFisher, USA); extracted using a custom protocol [16] on a Hamilton Microlab STAR (Hamilton, USA) and tested using the AccuPower SARS-CoV-2 Real-Time RT-PCR Kit (Bioneer, Korea) on a ABI 7500 FAST; or loaded directly to a Roche cobas® 6800 system and assayed with the cobas® SARS-CoV-2 Test (Roche, Switzerland).

All laboratory testing was conducted at HMC Central Laboratory following standardized protocols.

### Statistical analysis

Frequency distributions were used to describe CMWs’ characteristics and to estimate different SARS-CoV-2 epidemiological measures. Anti-SARS-CoV-2 pooled mean prevalence across CMW communities was estimated using meta-analysis. Here, a DerSimonian-Laird random-effects model [17] was applied to pool seroprevalence measures that were weighted using the inverse-variance method [18, 19].

Chi-square tests and univariable logistic regressions were implemented to explore associations with anti-SARS-CoV-2 positivity. Odds ratios (ORs), 95% confidence intervals (CIs), and p-values were generated. Covariates with p-value ≤ 0.2 in univariable regression analysis were included in the multivariable logistic regression model where applicable. Covariates with p-value ≤ 0.05 in the multivariable model were considered as showing evidence for an association with the outcome. The distribution of PCR cycle threshold (Ct) values among persons testing PCR positive was further generated, and summary statistics reported.

## Results

A total of 4,970 CMWs from the ten CMW communities participated in this study (Table 1). Participants were mostly men (95.0%), below 40 years of age (71.5%), and of Nepalese (43.0%), Indian (33.1%), or Bangladeshi (11.6%) origin. Regression analyses identified each of sex, nationality, and CMW Community to be independently associated with seropositivity.

**Table 1.**
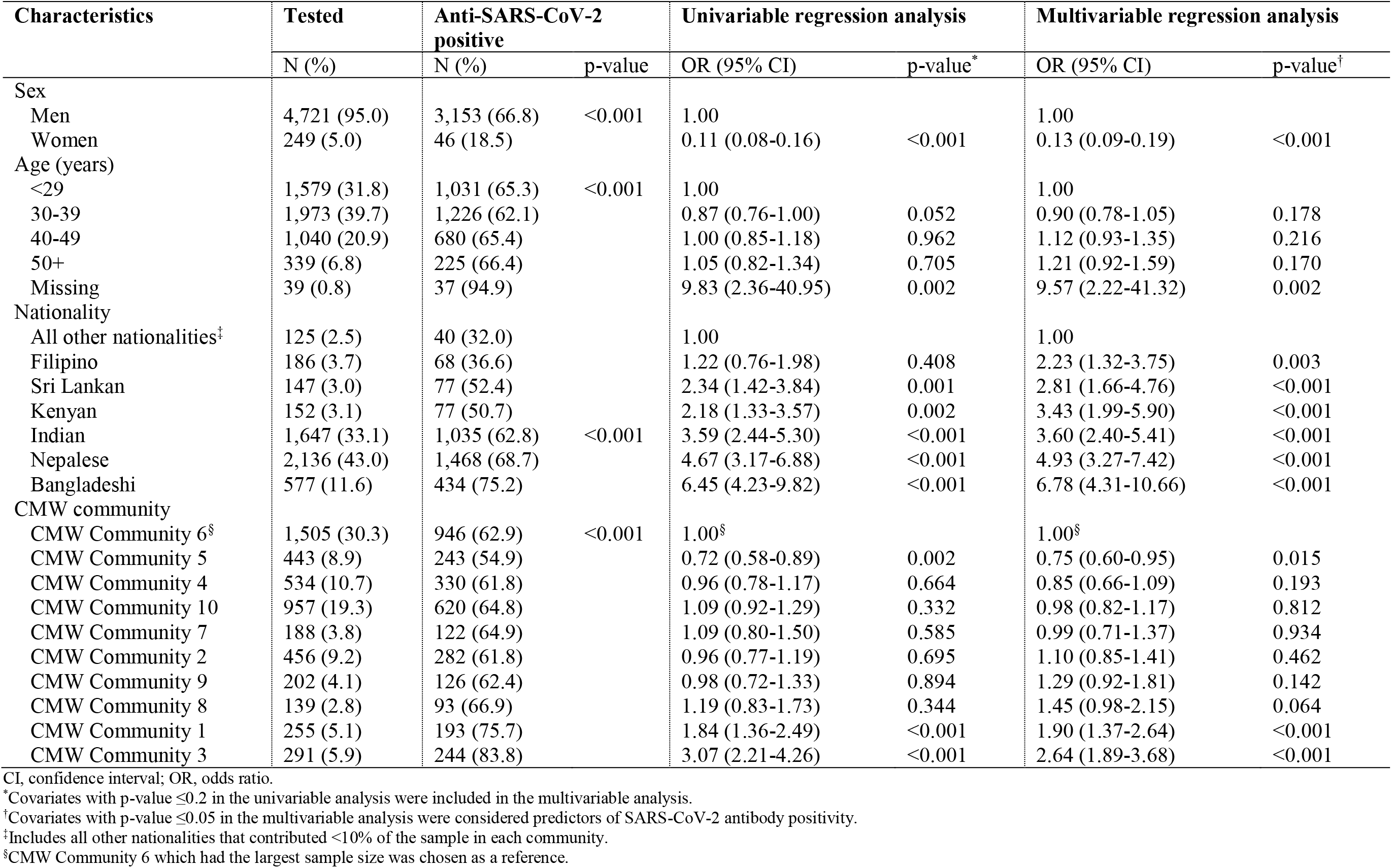
Characteristics of the craft and manual workers (CMWs) and associations with anti-SARS-CoV-2 positivity.

Women had 87% lower odds of being seropositive compared to men (adjusted odds ratio (AOR): 0.13; 95% CI: 0.09-0.19; Table 1). Compared to all other nationalities, AOR was 6.78 (95% CI: 4.31-10.66) for Bangladeshis, 4.93 (95% CI: 3.27-7.42) for Nepalese, 3.60 (95% CI: 2.40-5.41) for Indians, 3.43 (95% CI: 1.99-5.90) for Kenyans, 2.81 (95% CI: 1.66-4.76) for Sri Lankans, and 2.23 (95% CI: 1.32-3.75) for Filipinos. Some differences in seropositivity by CMW Community were noted. No significant differences in seropositivity by age group were found.

Table 2 shows the characteristics and associations with anti-SARS-CoV-2 positivity for only CMW Community 1 where a specific interview schedule was administered and collected specific socio-demographic data and history of exposure and symptoms. Close to 40% of participants had intermediate or low educational attainment, and a third had higher schooling levels or vocational training. University education was associated with 75% (OR: 0.25; 95% CI: 0.09-0.67) lower odds of seropositivity compared to intermediate or lower educational attainment. No statistically-significant associations with seropositivity were found for contact with an infected person, presence of symptoms, or symptoms requiring medical attention. Appendix Table S1 shows also the characteristics and associations with anti-SARS-CoV-2 positivity for CMW Communities 2-10. For each of these communities, associations were found for sex and nationality, but no notable associations were found for age group.

**Table 2.**
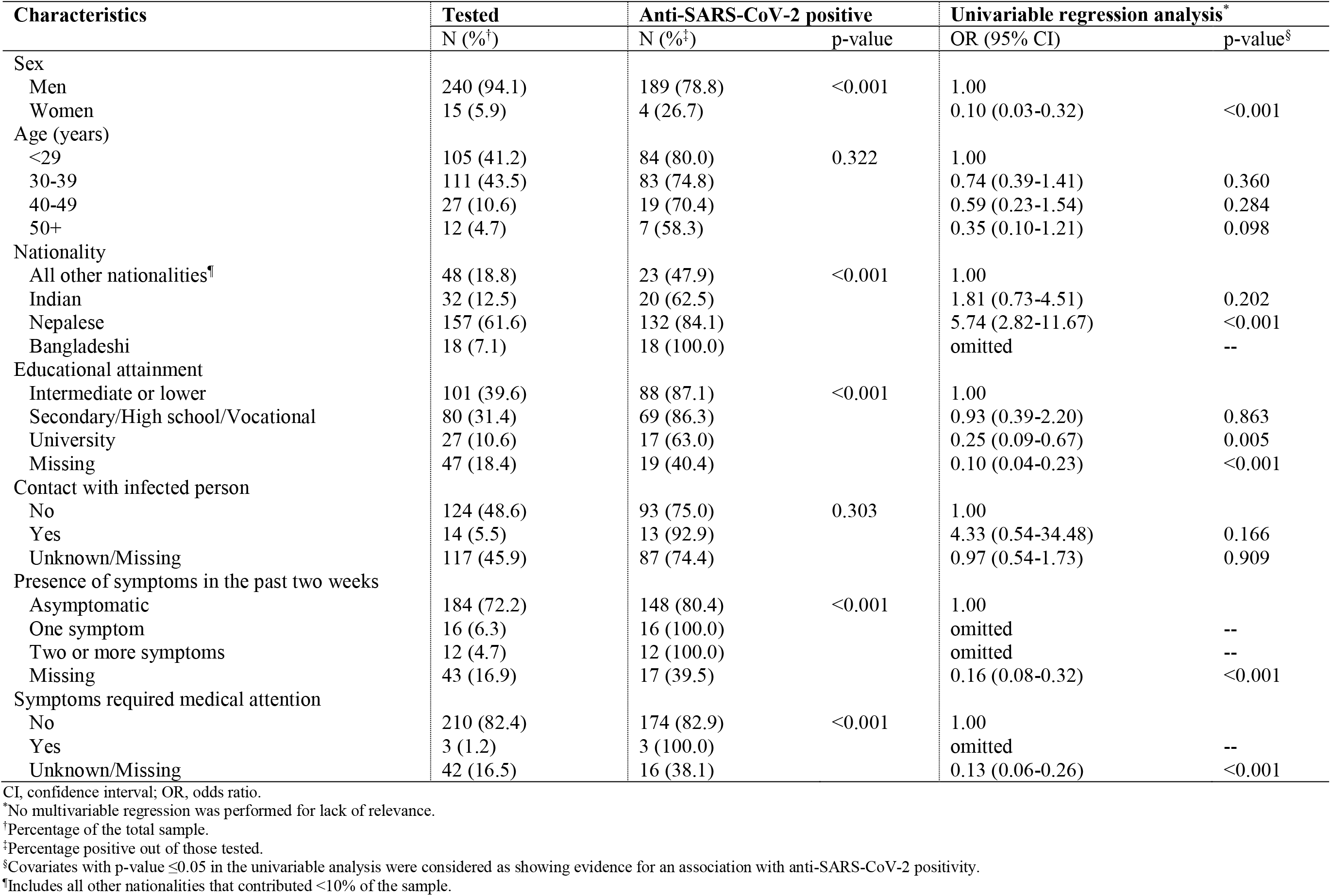
Characteristics of the Craft and Manual Worker (CMW) Community 1 and associations with anti-SARS-CoV-2 positivity including socio-demographics and history of exposure and symptoms.

Figure 1 illustrates key SARS-CoV-2 epidemiological measures in the different CMW communities. Out of a total of 4,970 anti-SARS-CoV-2 test results for these CMWs, 3,199 (64.4%; 95% CI: 63.0-65.7%) were seropositive. Seropositivity ranged from 54.9% (95% CI: 50.2-59.4%) in CMW Community 5 to 83.8% (95% CI: 79.1-87.7%) in CMW Community 3 (Figure 1A). The pooled mean anti-SARS-CoV-2 positivity across the ten CMW communities was 66.1% (95% CI: 61.5-70.6%).

**Figure 1.**
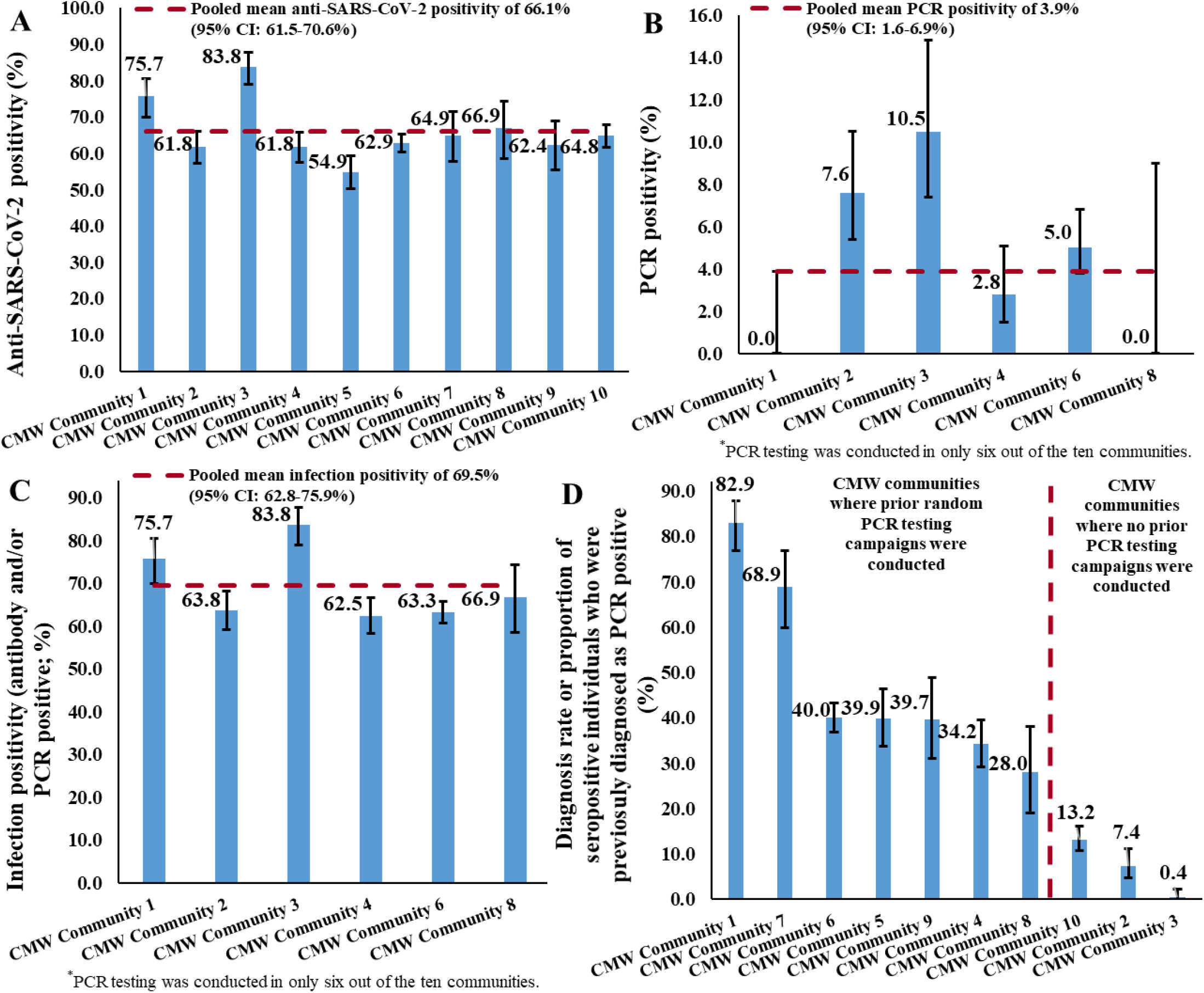
Measures of SARS-CoV-2 A) antibody positivity, B) PCR positivity, C) infection positivity (antibody and/or PCR positive), and D) diagnosis rate, across the craft and manual worker (CMW) communities. Of note that PCR testing was done in only six communities.

Out of a total of 2,016 PCR test results for these CMWs, 112 (5.6%; 95% CI: 4.6-6.6%) were positive. PCR positivity ranged from 0.0% (95% CI: 0.0-3.9%) in CMW Community 1 and 0.0% (95% CI: 0.0-9.0%) in CMW Community 8 to 10.5% (95% CI: 7.4-14.8%) in CMW Community 3 (Figure 1B). Pooled mean PCR positivity across the six CMW communities where PCR testing was conducted was 3.9% (95% CI: 1.6-6.9%). Ct values ranged from 15.8-37.4 with a median of 34.0 (Figure 2). The vast majority (79.5%) of PCR-positive individuals had a Ct value >30 suggestive of no active infection [20, 21]. Significant differences in PCR positivity were found by nationality and CMW Community (Appendix Table S2).

**Figure 2.**
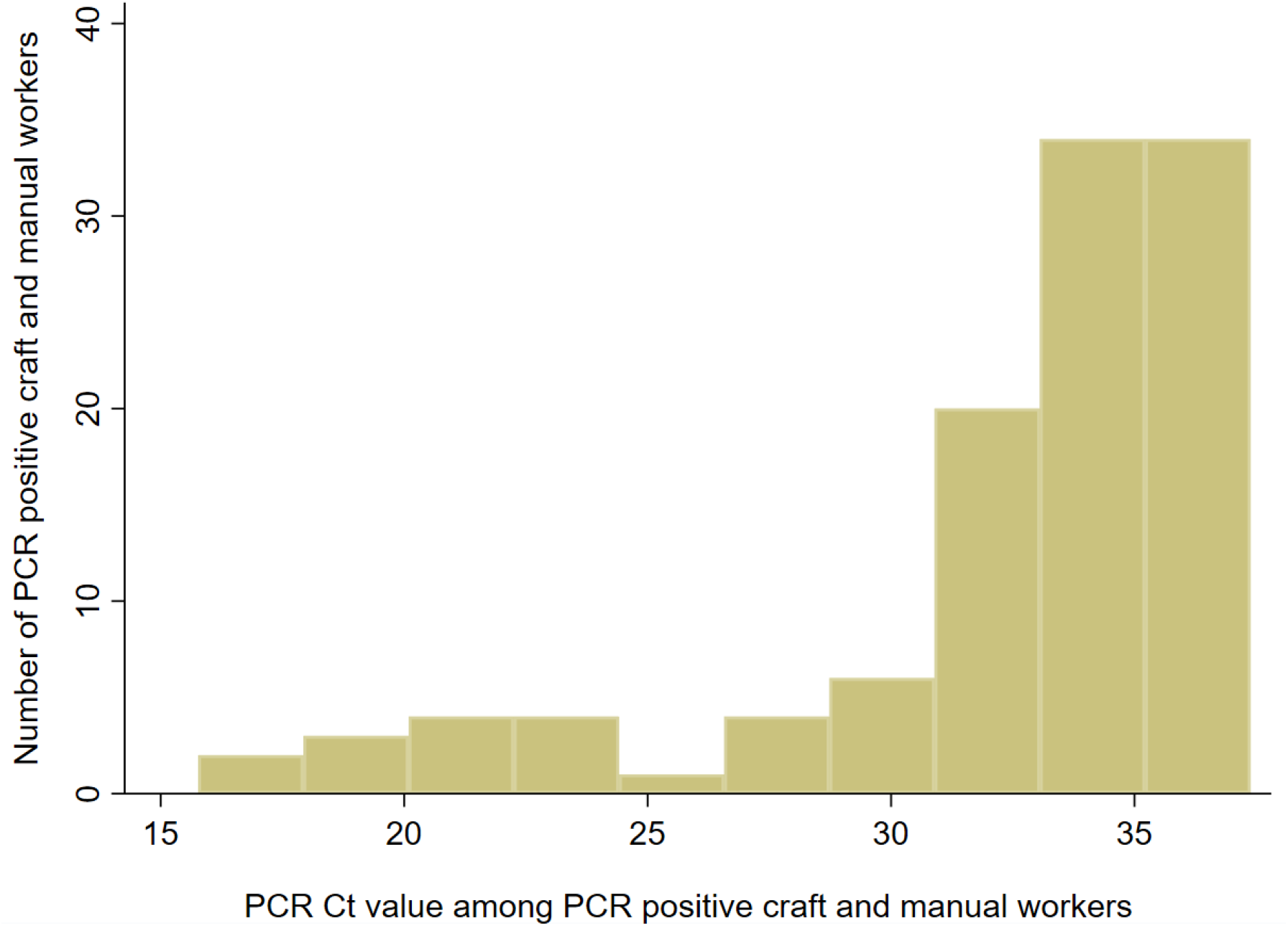
Distribution of PCR cycle threshold (Ct) values among craft and manual workers (CMWs) identified as SARS-CoV-2 PCR positive during the study period.

Infection positivity (antibody and/or PCR positive) ranged from 62.5% (95% CI: 58.3-66.7%) in CMW Community 4 to 83.8% (95% CI: 79.1-87.7%) in CMW Community 3 (Figure 1C). Pooled mean infection positivity across the six CMW communities with antibody and PCR results was 69.5% (95% CI: 62.8-75.9%).

Data were linked to the national SARS-CoV-2 PCR testing and hospitalization database. Of the 3,199 antibody positive CMWs, 1,012 (31.6%; 95% CI: 30.0-33.3%) were *previously* diagnosed with SARS-CoV-2 infection (had a laboratory-confirmed PCR positive result *before* this study). For the CMW communities that were previously part of broad PCR testing because of a case identification and/or a random testing campaign, the diagnosis rate ranged from 28.0% (95% CI: 19.1-38.2%) in CMW Community 8 to 82.9% (95% CI: 76.8-87.9%) in CMW Community 1.

Meanwhile, where no such broad PCR testing was conducted, the diagnosis rate was only 13.2% (95% CI: 10.7-16.1%) in CMW Community 10, 7.4% (95% CI: 4.7-11.2%) in CMW Community 2, and 0.4% (95% CI: 0.0-2.3%) in CMW Community 3. Only a very small fraction of antibody *negative* persons, 14 out of 1,771 (0.8%; 95% CI: 0.4-1.3%), had been previously diagnosed as PCR positive (Appendix Table S3).

Of the total sample, 21 individuals had a hospitalization record associated with a SARS-CoV-2 infection diagnosis, of whom, infection severity per WHO classification was mild for five, moderate for ten, severe for five, and critical for one. All 21 individuals eventually cleared their infection and were discharged from the hospital. All these individuals also tested anti-SARS-CoV-2 positive. Accordingly, the proportion of those with a confirmed *severe* or *critical* infection out of 3,233 who had a laboratory-confirmed infection (antibody and/or PCR positive result) was 0.2% (95% CI: 0.1-0.4%).

## Discussion

Above results support that herd immunity has been reached (or at least nearly reached) in these CMW communities, and that the level of herd immunity needed for SARS-CoV-2 infection is an attack rate (proportion ever infected) of about 65-70%.

This conclusion is supported by: i) these CMW communities had comparable seroprevalence of about 65-70%; ii) PCR positivity was low and the vast majority of those PCR positive had high Ct value suggestive of an earlier rather than recent infection [20, 21]; iii) only few persons had active infection (Ct value <25) and no significant infection cluster was identified in any of these CMW communities during this study (suggestive of isolated infections and unsustainable infection transmission for clusters to occur); iv) the level of 65-70% infection exposure is in concordance with that predicted using the “classical” formula for herd immunity of ∼1−1 *R*_0_ [22, 23], with *R*_*0*_, the basic reproduction number, being at 2.5-4 [24, 25].

Notably, although large clusters of infection were very common in such CMW communities before and around the epidemic peak towards end of May, no major cluster has been identified in any CMW community in Qatar for several weeks now, despite the progressive easing of the social and physical distancing restrictions since June 15, 2020.

These findings indicate that reaching herd immunity in such largely homogenous communities requires high exposure level of about 65-70%. However, it is possible that *true* herd immunity may have been reached even at a lower attack rate. Mathematical modeling indicates that infection exposure for a novel infection (especially in the first cycle) can considerably “overshoot” the “classical” herd immunity level of ∼1−1 *R*_0_, more so if the social contact rate within this community is homogeneous (illustration is shown in Appendix Figure S1). Meanwhile, heterogeneity in social contact rate can reduce the final attack rate (Appendix Figure S1) [23, 26].

This study had other notable findings. Severity rate for SARS-CoV-2 infection was low (0.2%), possibly because of the young age of the CMWs. No COVID-19 deaths were reported in these CMW communities. Remarkably, in the communities where no prior broad PCR testing was conducted, <15% of the antibody positive subjects had ever been diagnosed as PCR positive prior to this study. There was a large difference in infection exposure between women and men (Table 1). This difference, with the variable proportion of women across these communities, explains also part of the variation seen in seroprevalence across these communities (Figure 1 versus Appendix Figure S2). This finding may be attributed to women and men living in different housing accommodations and having different work roles. Women, a very small minority in these CMW communities, live in small shared accommodations as opposed to the large ones hosting men.

There were differences by nationality (Table 1), but these are explained by nearly all Bangladeshis and Nepalese and most Indians being the workers in these communities, while a proportion of Indians and much of the other nationalities holding administrative or managerial positions with lower social contact rates and possibly living in a different kind of accommodations compared to the bulk of the workers. No major differences in infection exposure by age were found, though there was some tendency for those >40 years of age to have lower infection exposure (Appendix Table S1), possibly due to different occupations within these communities.

This study has limitations. Testing was conducted in select CMW communities and therefore findings may not be generalizable to the wider CMW population in Qatar. Response rate could not be precisely ascertained given uncertainty around the number of CMWs who were aware of the invitation to participate, but based on employer-reported counts of the size of each community, the response rate is >50% and participants expressed high interest in knowing their antibody status. The validity of study outcomes is contingent on the sensitivity and specificity of the used assays. However, the laboratory methods were based on high-quality commercial platforms, and each diagnostic method was validated in the laboratory before its use. Notably, the antibody assay had high specificity reported at 99.8% [15] by the manufacturer and at 100% by a validation study by Public Health England [27].

In conclusion, at least some of the CMW communities in Qatar, who constitute about 60% of the total population [8], have reached or nearly reached herd immunity for SARS-CoV-2 infection, providing to our knowledge the first empirical evidence for herd immunity worldwide. While achieving herd immunity at a national level is difficult within few months [28], herd immunity could be achieved in specific communities within few months. In such relatively homogenous communities, reaching herd immunity required infection of 65-70% of the members of the community. These findings suggest that the SARS-CoV-2 epidemic in a homogenous population is unlikely to be unsustainable before as much as two-thirds of the population become infected. This also suggests that a SARS-CoV-2 vaccine needs at least 65-70% efficacy at universal coverage for herd immunity to be achieved in a population naïve to SARS-CoV-2 infection [29].

## Data Availability

All data are available in an aggregate form in the main text and supplementary materials.

## Author contributions

AJ and LJA co-conceived and co-designed the study. AJ led data collection. HC performed the data analyses and wrote the first draft of the article. HHA contributed to analysis of data. All authors contributed to data acquisition, database development, testing, program development, discussion and interpretation of the results, and to the writing of the manuscript. All authors have read and approved the final manuscript.

## Acknowledgement

We would like to thank Her Excellency Dr. Hanan Al Kuwari, the Minister of Public Health, for her vision, guidance, leadership, and support. We also would like to thank Dr. Saad Al Kaabi, Chair of the System Wide Incident Command and Control (SWICC) Committee for the COVID-19 national healthcare response, for his leadership, analytical insights, and for his instrumental role in enacting the data information systems that made these studies possible. We further extend our appreciation to the SWICC Committee and the Scientific Reference and Research Taskforce (SRRT) members for their informative input, scientific technical advice, and enriching discussions. We would also like to thank Dr. Mariam Abdulmalik, the CEO of the Primary Health Care Corporation and the Chairperson of the Tactical Community Command Group on COVID-19, as well as members of this committee, for providing support to the teams that worked on the field surveillance. We also would like to acknowledge the dedicated efforts of the Clinical Coding Team and the COVID-19 Mortality Review Team, both at Hamad Medical Corporation, and the Surveillance Team at the Ministry of Public Health. We further would like to acknowledge Mr. Ziad Yehya, the COVID-19 Incident Commander and Corporate Planning and Enterprise Risk manager at Qatargas for dedicated logistical and resource support. We further thank all companies that have facilitated the participation of their employees in this study.

## Appendix

**Table S1.**
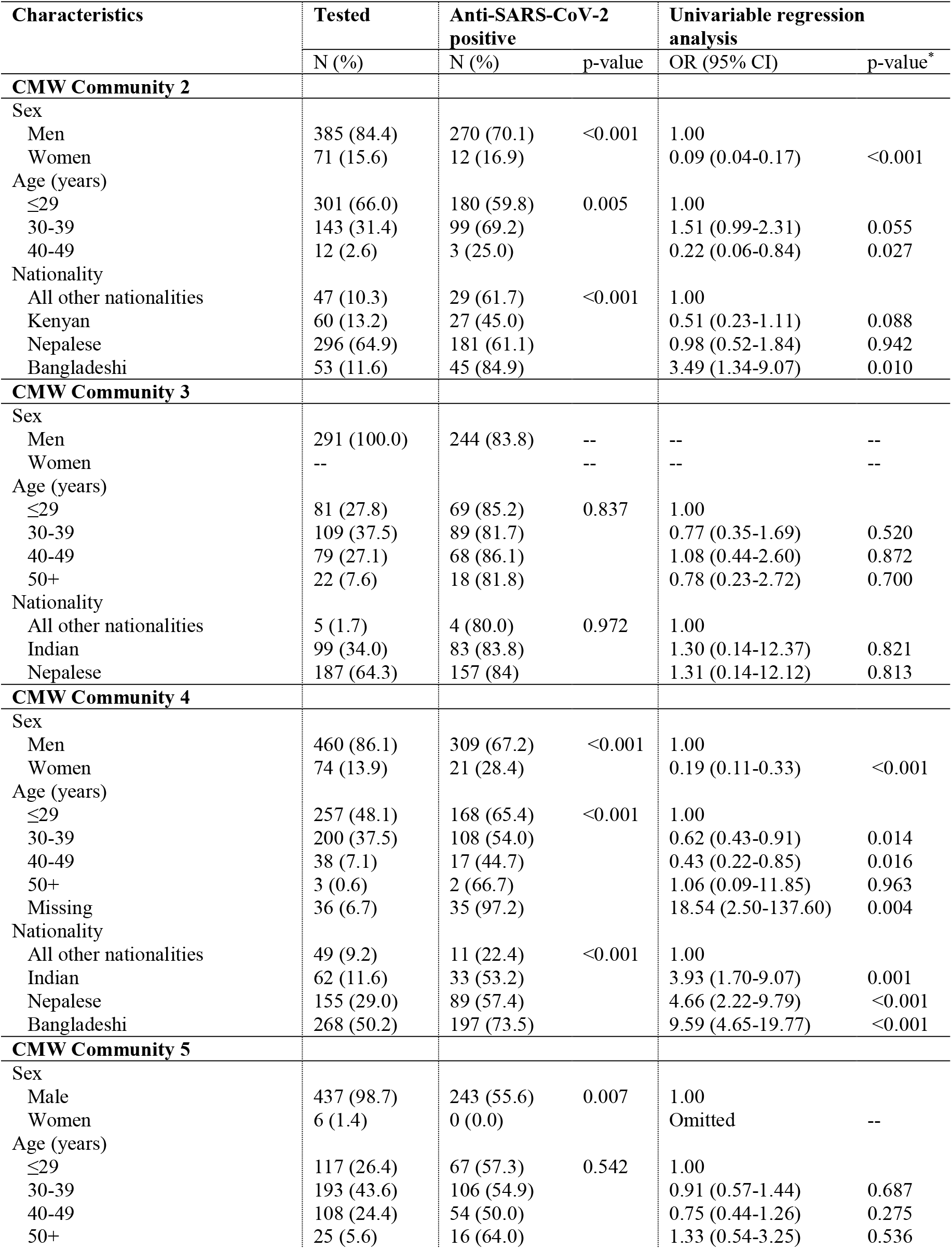
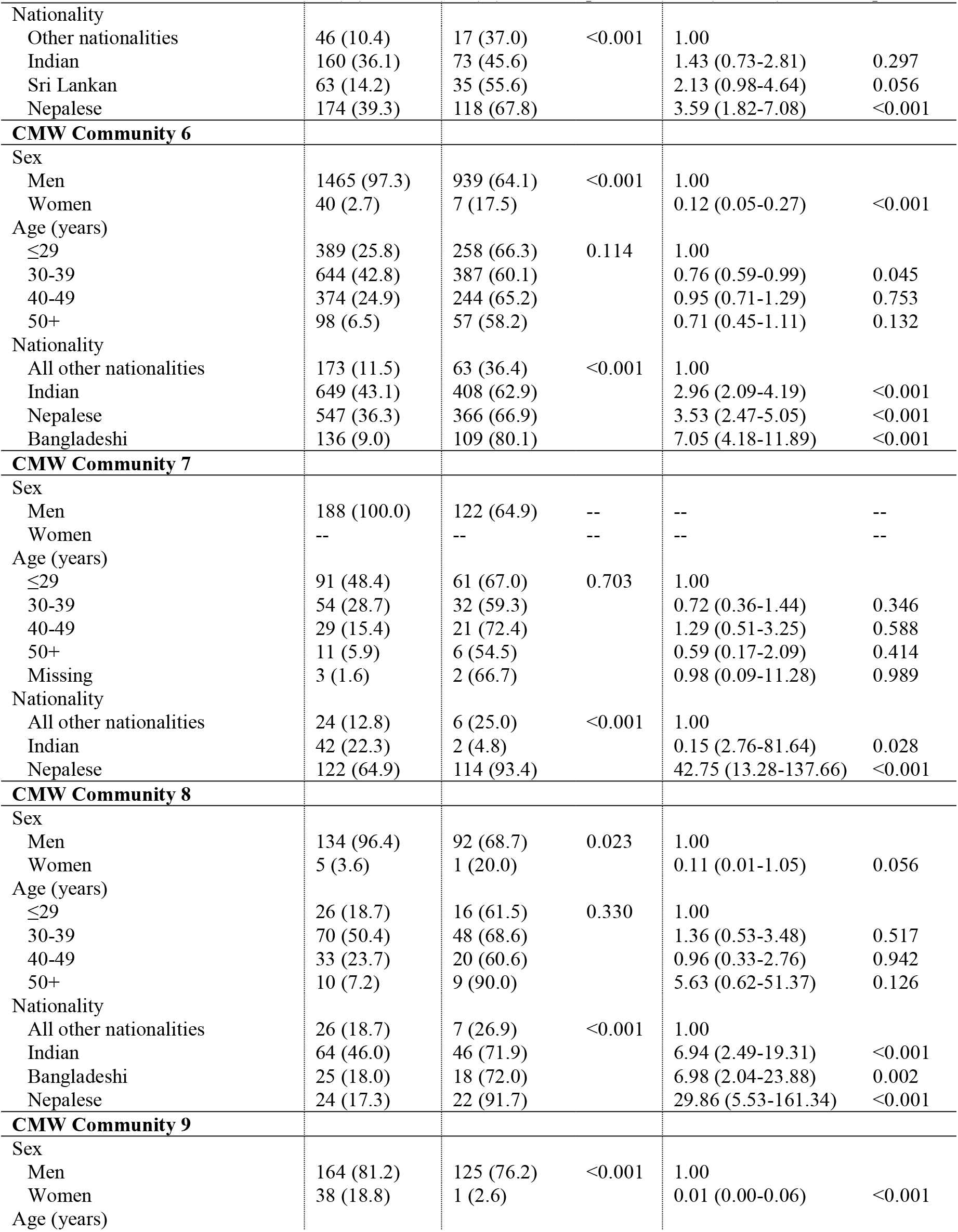
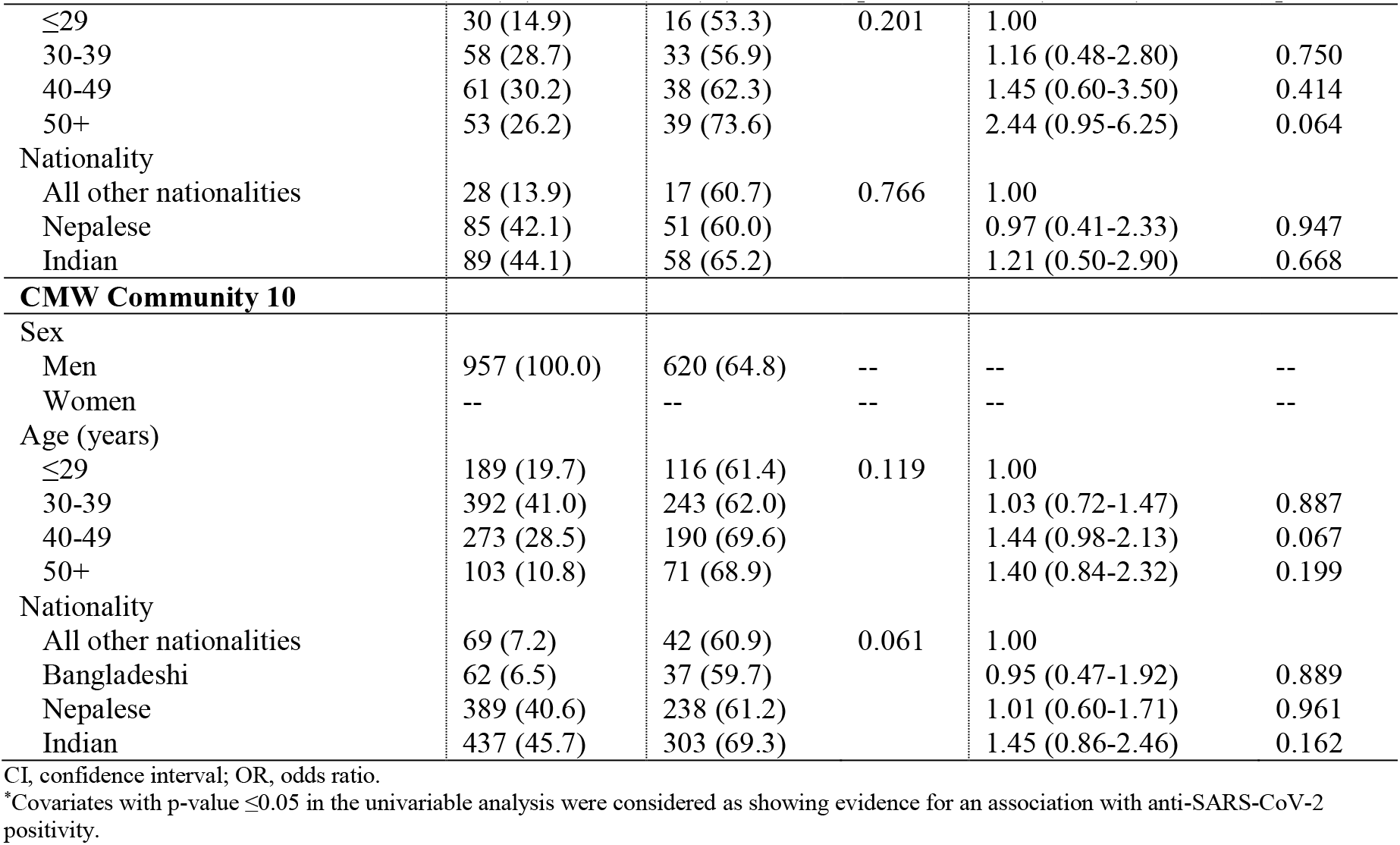
Characteristics of the Craft and Manual Worker (CMW) Communities 2-10 and associations with anti-SARS-CoV-2 positivity.

**Table S2.**
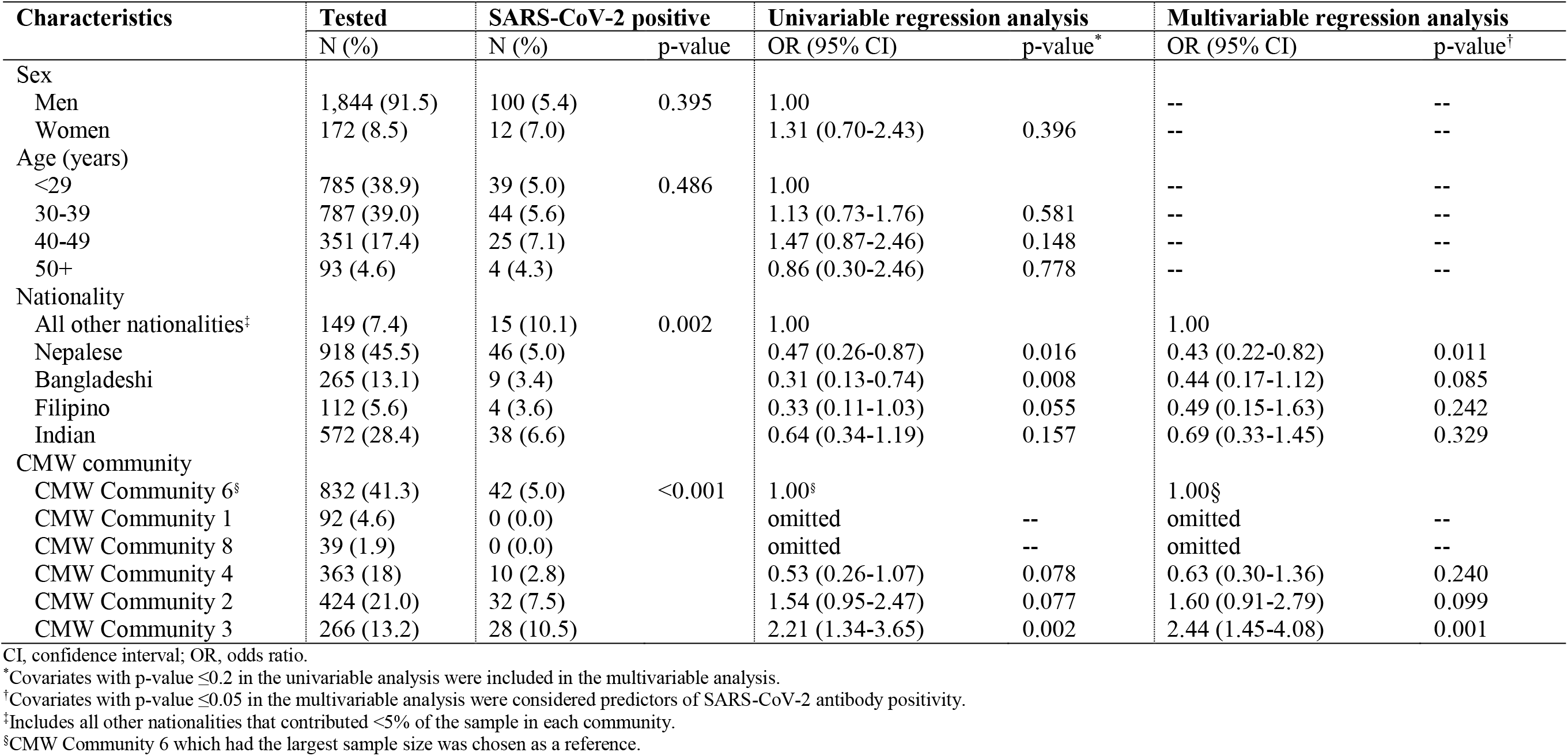
Characteristics of the craft and manual workers (CMWs) and associations with SARS-CoV-2 polymerase chain reaction (PCR) positivity.

**Table S3.** Characteristics of anti-SARS-CoV-2 negative persons who had tested positive for SARS-CoV-2 infection using polymerase chain reaction (PCR) at some point prior to conduct of this study.

| Person number | Age | Sex | PCR test date | Ct value | Antibody test date | Symptoms |
| --- | --- | --- | --- | --- | --- | --- |
| 1 | 36 | Man | 13 May | 35.5 | 02 August | Not recorded |
| 2 | 28 | Man | 04 May | 34.1 | 03 August | Not recorded |
| 3 | 25 | Man | 04 May | 35.1 | 03 August | Not recorded |
| 4 | 26 | Man | 28 April | 22.0 | 08 August | Not recorded |
| 5 | 40 | Man | 06 June | 35.5 | 23 July | Not recorded |
| 6 | 33 | Man | 09 June | 23.7 | 27 July | Not recorded |
| 7 | 27 | Man | 12 May | 35.8 | 28 July | Not recorded |
| 8 | 34 | Man | 13 May | Unknown | 28 July | Not recorded |
| 9 | 31 | Man | 23 May | 27.4 | 20 August | Not recorded |
| 10 | 31 | Man | 06 May | 19.6 | 20 August | Not recorded |
| 11 | 33 | Man | 07 May | 18.0 | 20 August | Not recorded |
| 12 | 39 | Man | 11 July | 32.9 | 20 August | Not recorded |
| 13 | 44 | Man | 05 August | 35.2 | 20 August | Asymptomatic |
| 14 | 34 | Man | 08 May | 32.0 | 24 August | Not recorded |

**Figure S1.**
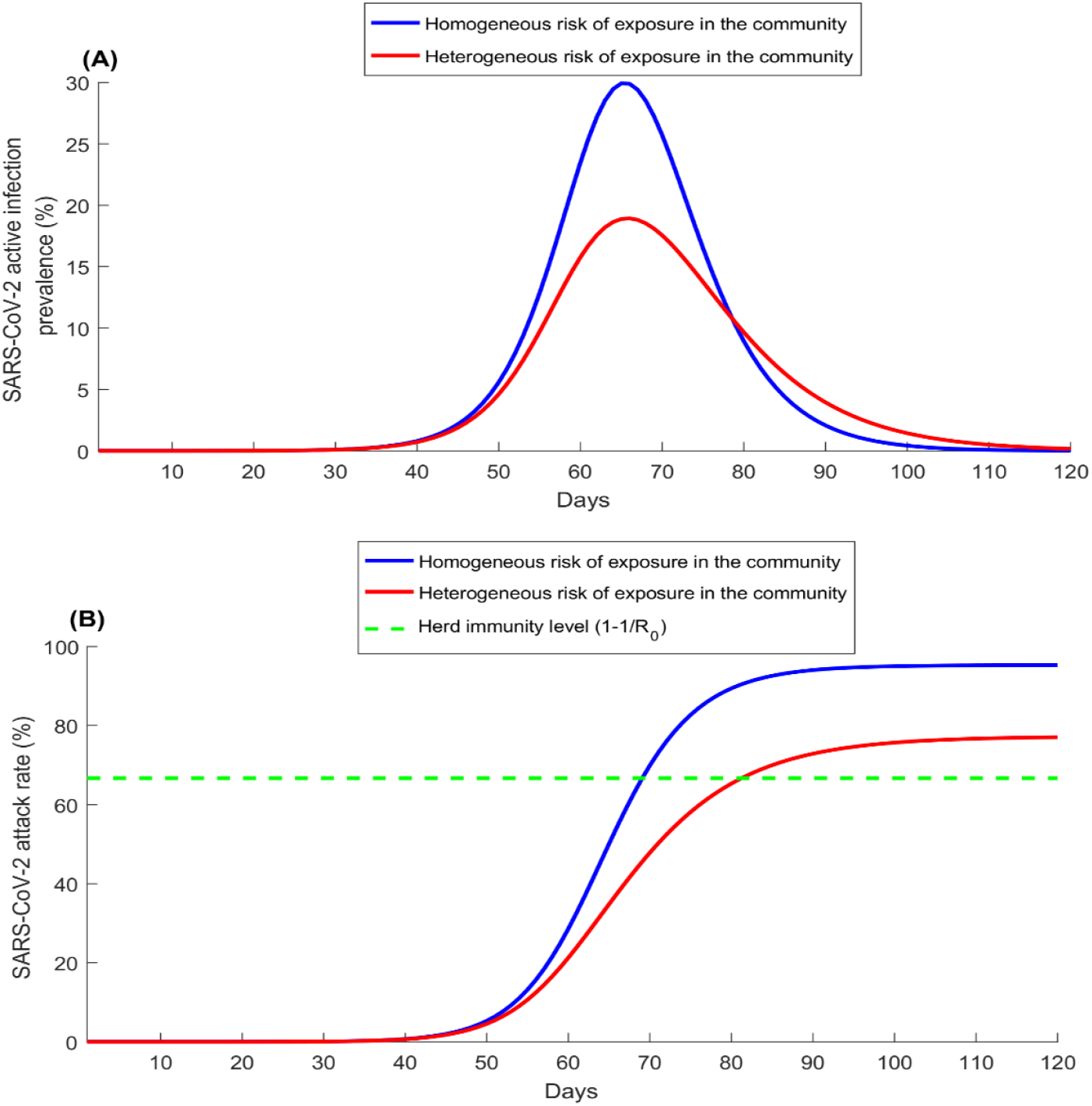
Herd immunity and heterogeneity in risk of exposure to the infection. SARS-CoV-2 active infection prevalence (A) and attack rate (proportion of the population that has ever been infected) (B) in a community where the risk of exposure is homogeneous versus in a community where the risk of exposure is heterogeneous. In both of these scenarios, the basic reproduction number *R*_0_ was assumed equal to 3 [1, 2]. These simulations were generated using a classic age-structured susceptible-exposed-infectious-recovered “SEIR” mathematical model [3]. Heterogeneity in the second modeled scenario was introduced through variable exposure risk by age.

**Figure S2.**
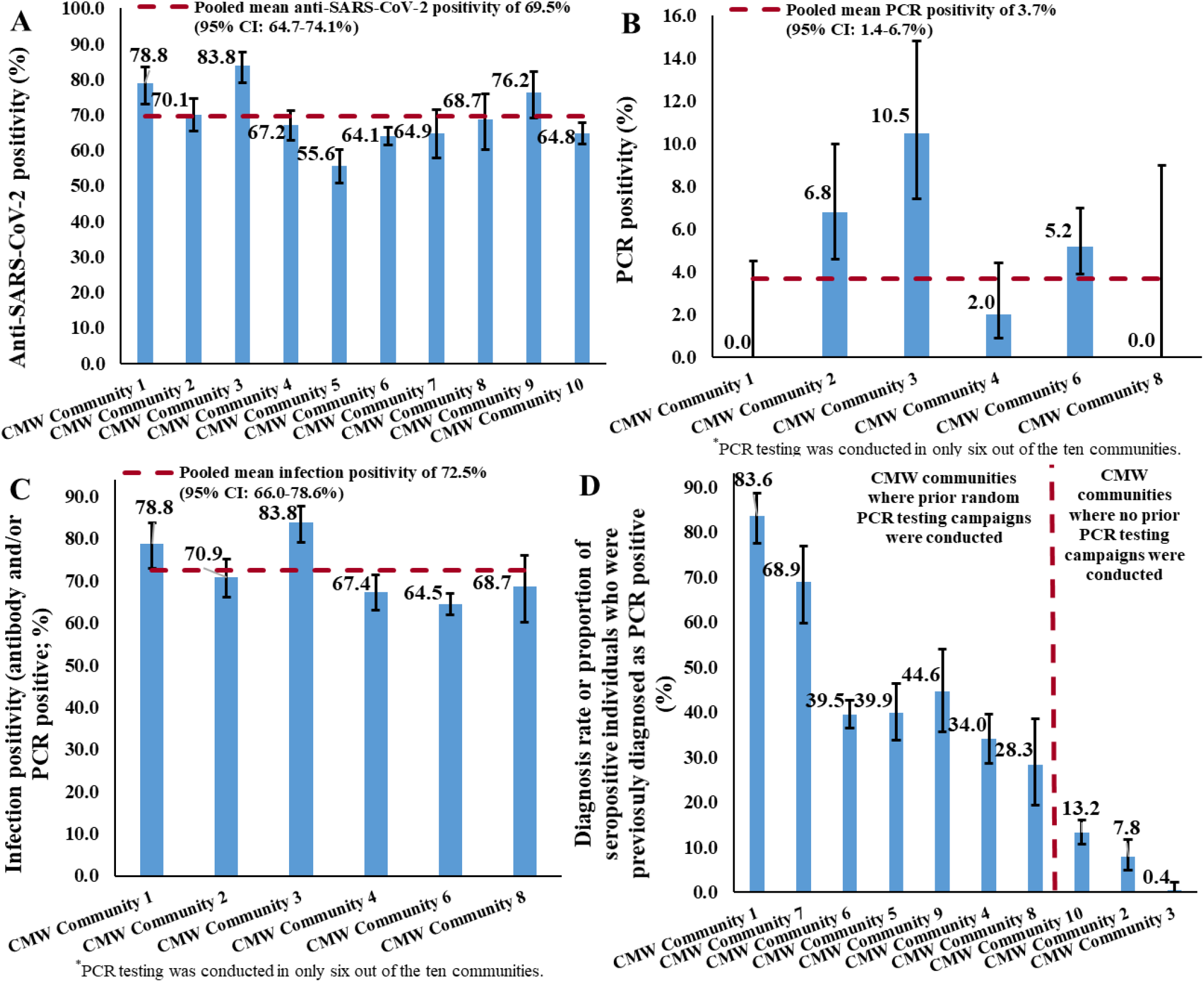
Measures of SARS-CoV-2 A) antibody positivity, B) PCR positivity, C) infection positivity (antibody and/or PCR positive), and D) diagnosis rate, among *only men* craft and manual workers (CMW) across the CMW communities.

## References

1. COVID-19 Outbreak Live Update. Available from: https://www.worldometers.info/coronavirus/. xAccessed on September 06, 2020. 2020.

2. Ioannidis, J., The infection fatality rate of COVID-19 inferred from seroprevalence data. medRxiv, 2020: p. 2020.05.13.20101253.

3. Planning and Statistics Authority-State of Qatar, Qatar Monthly Statistics. Available from: https://www.psa.gov.qa/en/pages/default.aspx. Accessed on: may 26,2020. 2020.

4. Abu-Raddad, L.J., et al., Characterizing the Qatar advanced-phase SARS-CoV-2 epidemic. medRxiv, 2020: p. 2020.07.16.20155317v2.

5. Al Kuwari, H.M., et al., Characterization of the SARS-CoV-2 outbreak in the State of Qatar, February 28-April 18, 2020. BMJ open (accepted for publication), 2020.

6. Hamad Medical Corporation, SARS-CoV-2 hospitalizations and care. 2020.

7. Ministry of Public Health-State of Qatar, Coronavirus Disease 2019 (COVID-19). Available from: https://covid19.moph.gov.qa/EN/Pages/default.aspx. Accessed on: May 25, 2020. 2020.

8. Planning and Statistics Authority-State of Qatar, Labor force sample survey. Available from: https://www.psa.gov.qa/en/statistics/Statistical%20Releases/Social/LaborForce/2017/statistical_analysis_labor_force_2017_En.pdf. Accessed on: May 01, 2020. 2017.

9. Ministry of Interior-State of Qatar, Population distribution by sex, age, and nationality: results of Kashef database. 2020.

10. De Bel-Air, F., Demography, Migration, and Labour Market in Qatar. Available from: https://www.researchgate.net/publication/323129801_Demography_Migration_and_Labour_Market_in_Qatar-_UPDATED_June_2017. Accessed on May 01, 2020. 2018, Gulf Labour Markets and Migration.

11. Jackson, C., et al., School closures and influenza: systematic review of epidemiological studies. BMJ Open, 2013. 3(2).

12. Glatman-Freedman, A., et al., Attack rates assessment of the 2009 pandemic H1N1 influenza A in children and their contacts: a systematic review and meta-analysis. PLoS One, 2012. 7(11): p. e50228.

13. World Health Organization, Population-based age-stratified seroepidemiological investigation protocol for COVID-19 virus infection. Available from: file:///C:/Users/hsc2001/Downloads/covid-19-seroepidemiological-investigation-protocol-v3.pdf. Accessed on: April 15, 2020. 2020.

14. World Health Organization, Clinical management of COVID-19. Available from: https://www.who.int/publications-detail/clinical-management-of-covid-19. Accessed on: May 31st 2020. 2020.

15. The Roche Group, Roche’s COVID-19 antibody test receives FDA Emergency Use Authorization and is available in markets accepting the CE mark. Available from: https://www.roche.com/media/releases/med-cor-2020-05-03.htm. Accessed on: June 5, 2020. 2020.

16. Kalikiri, M.K.R., et al., High-throughput extraction of SARS-CoV-2 RNA from nasopharyngeal swabs using solid-phase reverse immobilization beads. medRxiv, 2020: p. 2020.04.08.20055731.

17. DerSimonian, R. and N. Laird, Meta-analysis in clinical trials. Control Clin Trials, 1986. 7(3): p. 177–88.

18. Miller, J.J., The Inverse of the Freeman – Tukey Double Arcsine Transformation. The American Statistician, 1978. 32(4): p. 138–138.

19. Barendregt, J.J., et al., Meta-analysis of prevalence. J Epidemiol Community Health, 2013. 67(11): p. 974–8.

20. Sethuraman, N., S.S. Jeremiah, and A. Ryo, Interpreting Diagnostic Tests for SARS-CoV-2. JAMA, 2020.

21. Wajnberg, A., et al., Humoral immune response and prolonged PCR positivity in a cohort of 1343 SARS-CoV 2 patients in the New York City region. medRxiv, 2020: p. 2020.04.30.20085613.

22. Anderson, R.M., et al., How will country-based mitigation measures influence the course of the COVID-19 epidemic? Lancet, 2020. 395(10228): p. 931–934.

23. Britton, T., F. Ball, and P. Trapman, A mathematical model reveals the influence of population heterogeneity on herd immunity to SARS-CoV-2. Science, 2020. 369(6505): p. 846–849.

24. He, W., G.Y. Yi, and Y. Zhu, Estimation of the basic reproduction number, average incubation time, asymptomatic infection rate, and case fatality rate for COVID-19: Meta-analysis and sensitivity analysis. J Med Virol, 2020.

25. MIDAS Online COVID-19 Portal, COVID-19 parameter estimates: basic reproduction number. Available from: https://github.com/midas-network/COVID-19/tree/master/parameter_estimates/2019_novel_coronavirus. Accessed on: MAy 19, 2020. 2020.

26. Aguas, R., et al., Herd immunity thresholds for SARS-CoV-2 estimated from unfolding epidemics. medRxiv, 2020: p. 2020.07.23.20160762.

27. Public Health England, Evaluation of Roche Elecsys AntiSARS-CoV-2 serology assay for the detection of anti-SARS-CoV-2 antibodies. Available from: https://assets.publishing.service.gov.uk/government/uploads/system/uploads/attachment_data/file/891598/Evaluation_of_Roche_Elecsys_anti_SARS_CoV_2_PHE_200610_v8.1_FINAL.pdf. Accessed on June 5, 2020. 2020.

28. Pollan, M., et al., Prevalence of SARS-CoV-2 in Spain (ENE-COVID): a nationwide, population-based seroepidemiological study. Lancet, 2020. 396(10250): p. 535–544.

29. Makhoul, M., et al., Epidemiological impact of SARS-CoV-2 vaccination: mathematical modeling analyses. medRxiv, 2020: p. 2020.04.19.20070805.

## References

1. He, W., G.Y. Yi, and Y. Zhu, Estimation of the basic reproduction number, average incubation time, asymptomatic infection rate, and case fatality rate for COVID-19: Meta-analysis and sensitivity analysis. J Med Virol, 2020.

2. MIDAS Online COVID-19 Portal, COVID-19 parameter estimates: basic reproduction number. Available from: https://github.com/midas-network/COVID-19/tree/master/parameter_estimates/2019_novel_coronavirus. Accessed on: MAy 19, 2020. 2020.

3. Abu-Raddad, L.J., et al., Characterizing the Qatar advanced-phase SARS-CoV-2 epidemic. medRxiv, 2020: p. 2020.07.16.20155317v2.

